# Rapid and sustained resolution of scrotal content pain through multimodal integrative neuromuscular interventions

**DOI:** 10.1101/2025.10.20.25338413

**Authors:** Christopher L Mendias, Tariq M Awan

**Affiliations:** Performance Medicine Institute, Phoenix, AZ, 85012, USA

**Keywords:** Orchialgia, Scrotal Pain, Scrotal Content Pain, Pain Management, Pelvic Floor Therapy, Exercise Therapy, Neuromuscular Re-Education

## Abstract

**Background:** Scrotal content pain (SCP) is a common and significant clinical challenge that significantly impairs quality of life. SCP appears to be caused by mechanical dysfunction of the lower abdominal wall, inguinal canal, or pelvic floor, which can lead to compressive neuropathy of the ilioinguinal and genitofemoral nerves creating a persistent pain-spasm cycle. Previous interventions have typically focused on either pain or muscle dysfunction individually, rather than using an integrative approach to address both aspects.

**Aims:** To report outcomes of a novel, multimodal, integrative neuromuscular protocol for men with SCP in a retrospective case series.

**Methods:** Twenty eight men (mean age 42.1±14.9 years) with chronic SCP underwent four weekly treatment sessions of a multimodal protocol consisting of class IV laser therapy, dry needling, pulsed electromagnetic field therapy, manual therapy, progressive therapeutic exercises, and oral tadalafil 5mg. Pain was measured using a Visual Analog Scale (VAS) before and after each session. Physical and mental health were assessed using the NIH PROMIS Global-10.

**Results:** VAS pain decreased significantly from 8.0±0.6 cm at baseline to 3.0±1.1 cm after the first session. By the end of the 4-week protocol, VAS pain was 0.8±0.8 cm. The PROMIS Global-10 Physical Health score increased by 27%, and the Mental Health score increased by 9.4%. Pain and PROMIS score improvements exceeded established MCIDs.

**Conclusion:** Patients with SCP can achieve rapid, significant, and sustained pain relief and quality of life improvements using a multimodal therapeutic approach that is superior to monotherapies.

## Introduction

Scrotal content pain (SCP) represents a significant clinical challenge that impairs quality of life (QOL) [1]. SCP accounts for up to 5% of urology office visits, but often times urologists do not have readily available treatment options for these patients [1]. The scrotum and its contents receive sensory innervation from the genital branch of the genitofemoral nerve and the ilioinguinal nerve, which travel within or adjacent to the spermatic cord and inguinal canal. The spermatic cord is ensheathed within three fascial layers derived from the abdominal wall – this intimate anatomical relationship means that mechanical dysfunction of the lower abdominal wall, inguinal canal, or pelvic floor can directly lead to nerve irritation or compression and referred pain in the scrotum.

Idiopathic SCP is increasingly thought to be neuromuscular in nature. Histology of spermatic cords from men with SCP identified a high prevalence of Wallerian degeneration in nerve fibers of the cremaster muscle and fascia, indicating a state of chronic nerve injury [2]. Myofascial restriction and hypertonicity within the inguinal canal can therefore create a compressive neuropathy of the ilioinguinal and genitofemoral nerves, leading to SCP. Activation of nociceptive neurons of the scrotum can lead to reciprocal activation of the motor neurons of the cremaster through interneuron intermediates in the spinal cord, resulting in a persistent pain-spasm cycle.

Class IV lasers are used extensively to treat pain by inhibiting TRPV channels present in Aδ and C nociceptive neurons [3]. This impairs the ability of nociceptive neurons to propagate signals, with no impact on motor neurons [3]. Lasers therefore have advantages over local anesthetics which broadly target both sensory and motor neurons – by acutely reducing pain while preserving motor function, lasers create an ideal therapeutic window to selectively block nociceptive pain and spasms, allowing patients to perform exercises and other therapeutic interventions that restore normal neuromuscular control and achieve a lasting resolution. Here we report outcomes of a novel, integrated protocol which used class IV lasers to temporarily reduce SCP without impacting motor neuron function, allowing for subsequent physiotherapy interventions that provided lasting treatment for men with SCP.

## Methods

### Participants and Procedures

This study includes 28 male patients (mean age 42.1±14.9 years) who were treated at our clinic. Prior to treatment, patients were evaluated by a urologist who ruled out pathologies such as hernia, tumor, or varicocele. Mean SCP symptom duration was 3.5±1.4 months. Urine leukocyte esterase or urine cultures confirmed the absence of infection. Prostatitis was considered unlikely, with PSA levels <4 ng/mL and no pain on digital rectal examination. Almost half of patients (46%) had been treated with antibiotics without benefit, even though there were no laboratory signs of infection. Nearly one-third (29%) of patients had a previous vasectomy, but this was performed >3 years before SCP symptoms developed.

Patients underwent four weekly treatment sessions (Figure 1A-B). The treatment combined five synergistic modalities: (i) class IV laser therapy (REMY Medical Laser ZFT-15FJ, Cheny Hill, NJ), applied from the inguinal canal origin to the distal scrotum; (ii) dry needling with needles (SEIRIN, Boston, MA) placed intradermally or subcutaneously over the inguinal canal; (iii) PEMF (BioElectronics, Frederick, MD) focused on the external abdominal ring and cremaster muscle; (iv) manual therapy consisting of myofascial release and proprioceptive neuromuscular facilitative stretching of the abdominopelvic and hip musculature; and (v) therapeutic exercises. Therapeutic exercises were selected based on the mobility and fitness level of the patient, starting with foundational exercises (isometric adduction, supine march, pelvic tilts, cat-cow, quadruped rocking, glute bridge), progressing to intermediate exercises (single leg bridge, double leg raise, step-downs, swiss ball rollouts, side-lying abduction, dead bugs, squats, Pallof press, body blade rotations) and then advanced exercises (Copenhagen plank, reverse lunges, reverse deadlifts, Bulgarian split-squats, Russian twists, bicycle crunches). Patients were instructed to perform exercises and stretching at home 2-3 times weekly, along with diaphragmatic breathing. Patients were also prescribed oral tadalafil 5 mg daily to promote pelvic blood flow [4]. To quantify the impact of treatment on pain and QOL, visual analog scale (VAS, 10 cm) pain was measured before and after each session, and the NIH PROMIS Global 10 Health Outcome Assessment [5] was measured prior to starting treatment and at the end of the final session.

**Figure 1.**
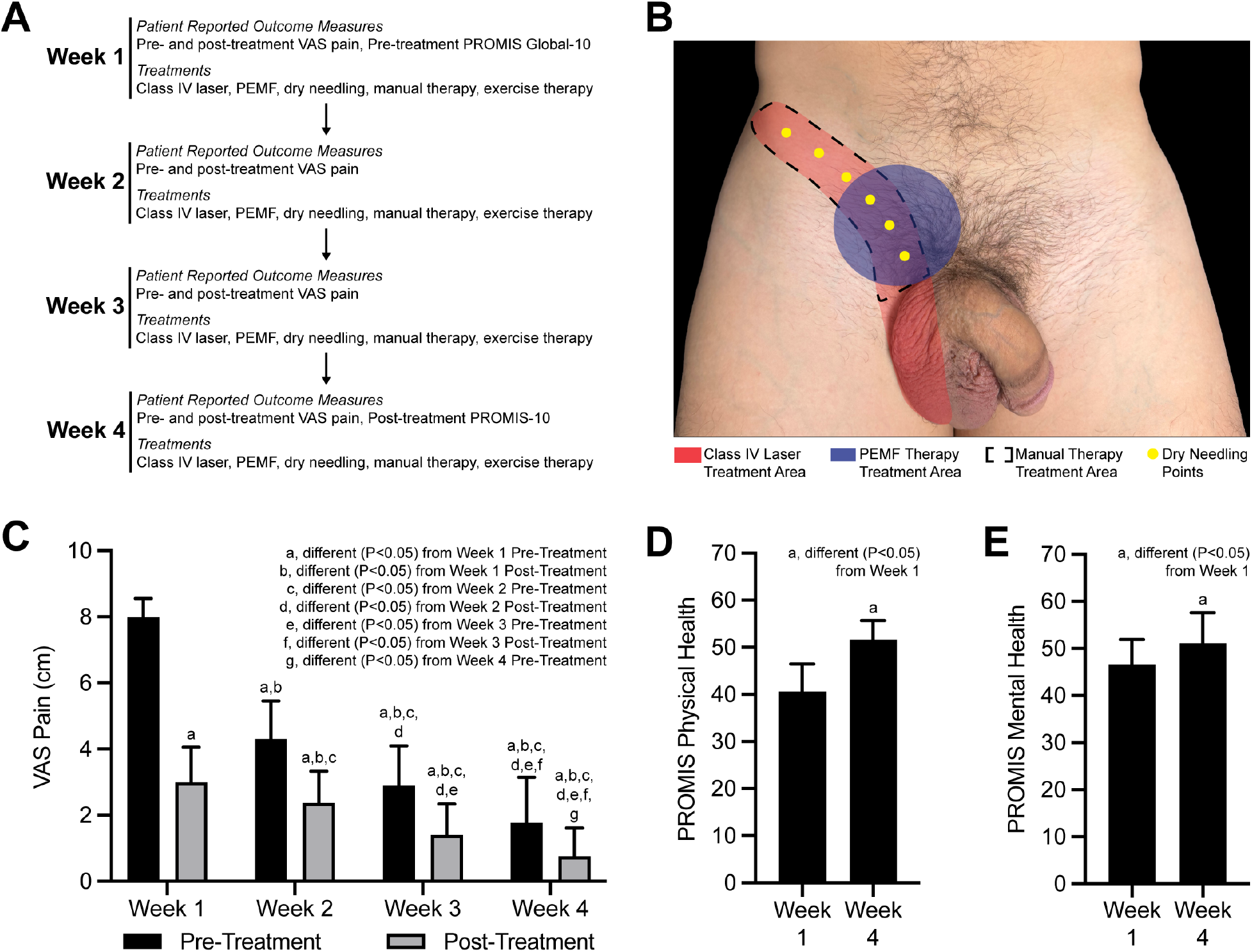
(A) Overview of timeline and study measures. (B) Representative treatment sites. (C) Changes in VAS Pain scores over time. Differences were tested with a two-way ANOVA followed by Tukey’s multiple comparisons test. PROMIS Global 10 (D) Physical Health and (E) Mental Health scores. Differences were tested with paired t-tests: a, significantly different (P<0.05) from Week 1. Values are mean±SD. N=28 male subjects. PEMF, Pulsed electromagnetic field therapy.

**Figure 2.**
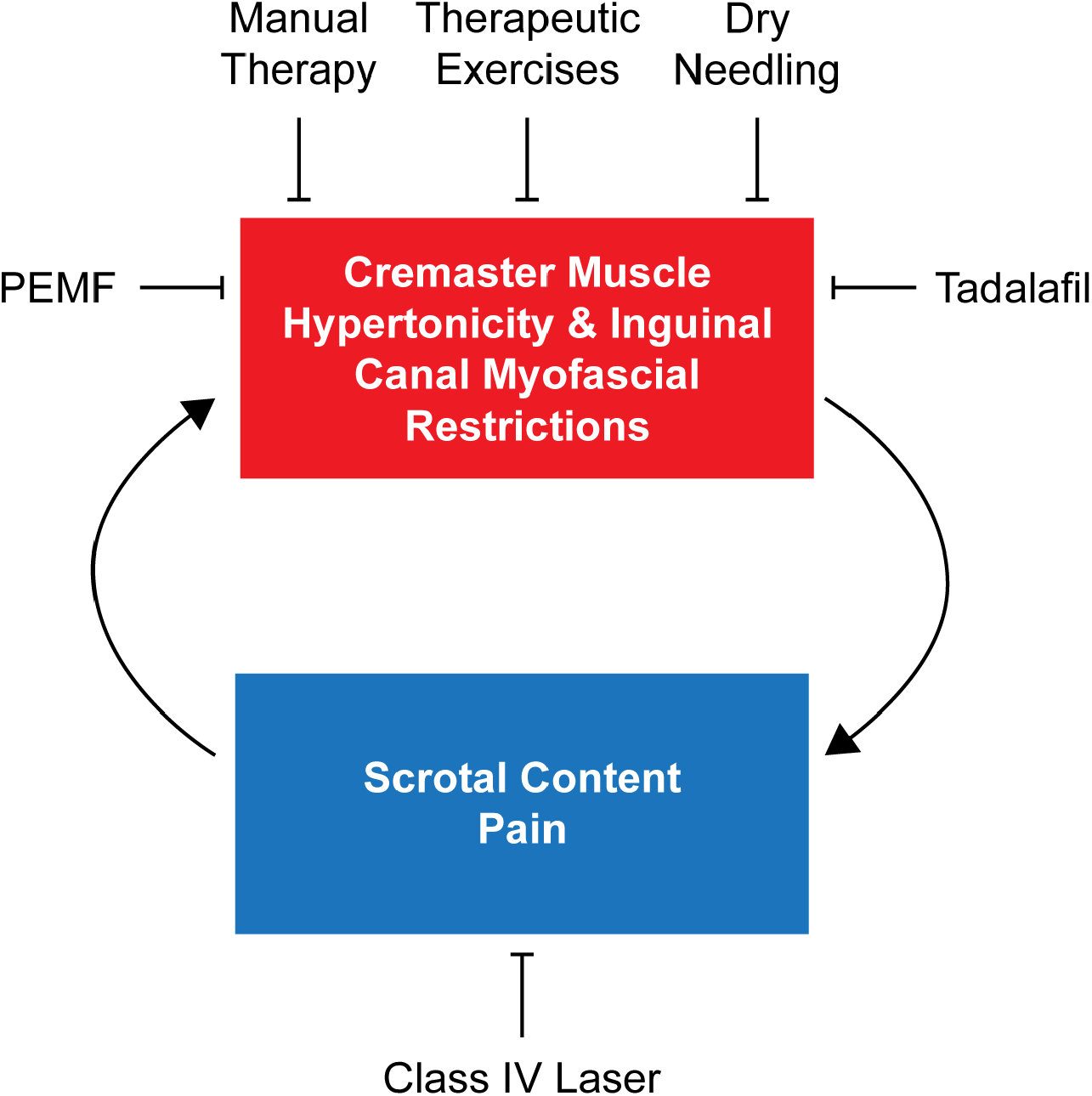
Proposed integrative model of treating scrotal content pain. PEMF, Pulsed electromagnetic field therapy.

### Statistics

Differences between pre- and post-treatment VAS pain scores over time were tested using a two-way ANOVA (α=0.05) followed by Tukey’s multiple comparisons tests. Paired t-tests (α=0.05) determined differences between Week 1 and Week 4 PROMIS Global 10 scores. Values are mean±SD.

## Results

Pain and QOL improved considerably over the 4-week program (Figure 1C-E). VAS pain scores decreased from 8.0±0.6 cm prior to starting treatment to 3.0±1.1 cm at the end of the first treatment session (P<0.05). Pain continued to decrease over 4 weeks, with a VAS pain of 0.8±0.8 cm (P<0.05) after the final session. This 90% reduction exceeded the minimal clinically important difference (MCID) of 30-50% for pelvic pain [6]. Physical and mental health also improved. The PROMIS Global-10 Physical Health score increased from 40.6±5.8 pre-treatment to 51.7±4.0 by Week 4, and the Mental Health score rose from 46.7±5.3 to 51.1±6.5, exceeding the MCID of 3.0 [5]. Follow-up at 3-6 months after therapy indicated that therapeutic gains were well maintained. Of the 25 (89%) who were successfully contacted, all reported treatment outcome satisfaction and denied recurrence of significant pain. No patients returned to the clinic for SCP-related issues during this follow-up period.

## Discussion

These findings demonstrate that a multimodal physiotherapy protocol can result in a rapid reduction in pain and an improvement in overall QOL. Our results support a neuromusculoskeletal etiology for many cases of SCP and highlight the efficacy of targeting the underlying mechanical and neurological dysfunctions using a polytherapeutic instead of monotherapeutic approach [7,8]. Each component of the intervention likely contributed synergistically to the overall outcomes. Laser therapy provided immediate pain modulation, creating a therapeutic window to apply other interventions [9]. Dry needling and PEMF addressed myofascial restrictions and pain. The combination of these modalities facilitated the subsequent application of manual therapy and therapeutic exercise, restoring normal neuromuscular function, improving mobility, and preventing recurrence of pain [10]. While this study is limited by its retrospective design, modest sample size, and the absence of a control group, the profound and consistent improvements observed in this cohort of patients are compelling.

## Conclusion

A multimodal physiotherapy protocol can provide rapid and sustained pain relief and QOL improvements for men with SCP. This study also supports the need for improved collaboration between urologists and musculoskeletal clinicians in the treatment of pelvic pain. Future prospective, randomized controlled trials are warranted to validate these findings and further establish the role of neuromuscular interventions in the treatment of SCP.

## Data Availability

All data produced in the present study are available upon reasonable request to the authors

## Acknowledgments

None

## References

[1] Sigalos JT, Pastuszak AW. Chronic orchialgia: epidemiology, diagnosis and evaluation. Transl Androl Urol 2017;6:S37–43. 10.21037/tau.2017.05.23.

[2] Parekattil SJ, Gudeloglu A, Brahmbhatt JV, Priola KB, Vieweg J, Allan RW. Trifecta Nerve Complex: Potential Anatomical Basis for Microsurgical Denervation of the Spermatic Cord for Chronic Orchialgia. J Urol 2013;190:265–70. 10.1016/j.juro.2013.01.045.

[3] Wang L, Zhang D, Schwarz W. TRPV Channels in Mast Cells as a Target for Low-Level-Laser Therapy. Cells 2014;3:662–73. 10.3390/cells3030662.

[4] Fukumoto K, Nagai A, Hara R, Fujii T, Miyaji Y. Tadalafil for male lower urinary tract symptoms improves endothelial function. Int J Urol 2017;24:206–10. 10.1111/iju.13273.

[5] Terwee CB, Peipert JD, Chapman R, Lai J-S, Terluin B, Cella D, et al. Minimal important change (MIC): a conceptual clarification and systematic review of MIC estimates of PROMIS measures. Qual Life Res 2021;30:2729–54. 10.1007/s11136-021-02925-y.

[6] Stephens-Shields AJ, Lai HH, Landis JR, Kreder K, Rodriguez LV, Naliboff BD, et al. Clinically Important Differences for Pain and Urinary Symptoms in Urological Chronic Pelvic Pain Syndrome: A MAPP Network Study. J Urol 2023;209:1132–40. 10.1097/ju.0000000000003394.

[7] Ergun O, Gudeloglu A, Parekattil SJ. Management of chronic orchialgia: review of current clinical practice. AME Méd J 2023;8:22–22. 10.21037/amj-22-98.

[8] Tawfik AM, Radwan MH, Abdulmonem M, Abo-Elenen M, Elgamal SA, Aboufarha MO. Tadalafil monotherapy in management of chronic prostatitis/chronic pelvic pain syndrome: a randomized double-blind placebo controlled clinical trial. World J Urol 2022;40:2505–11. 10.1007/s00345-022-04074-4.

[9] Momenzadeh C, ehghani-Ghorbi MD, Razzaghi MR, Abbasi MZ, Jaffari A. Influence of Low-Level Laser Irradiation of the Red and Infrared Spectral Range for Treating Chronic Testicular Pain: A Randomized Clinical Trial. J Lasers Méd Sci 2024;15:e62. 10.34172/jlms.2024.62.

[10] Corte-Rodriguez HDla, Roman-Belmonte JM, Resino-Luis C, Madrid-Gonzalez J, Rodriguez-Merchan EC. The Role of Physical Exercise in Chronic Musculoskeletal Pain: Best Medicine—A Narrative Review. Healthcare 2024;12:242. 10.3390/healthcare12020242.

